# Prognostic Significance of Left Ventricular Diastolic Dysfunction in the Community: A Project Baseline Health Study

**DOI:** 10.1101/2024.06.06.24308550

**Authors:** Gracia Fahed, Everton Santana, Nicholas Cauwenberghs, Bettia Celestin, Megan K. Carroll, arah A. Short, Kevin M. Alexander, Shadi P. Bagherzadeh, Kenneth W. Mahaffey, Adrian F. Hernandez, Svati H. Shah, Michael Salerno, Pamela S. Douglas, Tatiana Kusnetsova, Melissa A. Daubert, Francois Haddad, the Project Baseline Health Study Group

## Abstract

**Introduction:** Left ventricular diastolic dysfunction (LVDD) is an important predictor of cardiovascular outcomes. To date, however, few studies have validated the prognostic value of the LVDD assessment guidelines in a community-based setting. This study aimed to validate the predictive significance of LVDD following recommendations from the American Society of Echocardiography (ASE) and European Association of Cardiovascular Imaging (EACVI), and to explore the use of a diastolic score in characterizing LVDD, particularly in individuals who remain unclassified (indeterminate) by the guidelines.

**Materials and Methods:** Project Baseline Health Study (PBHS) participants were included (n=1,952). LVDD was graded using the ASE/EACVI criteria. Multivariable Cox regression estimated associations between LVDD and major adverse cardiovascular events (MACE). To gain insights into the indeterminate group, we developed a novel diastolic score (0-6 points) with minor and major criteria guided by percentile deviation from a healthy reference cohort (n=565), including early mitral annular tissue Doppler velocity (e’), the ratio of early diastolic inflow velocity to e’ (E/e’), and left atrial volume index.

**Results:** At enrollment, the mean age of participants was 50±16; and 56% were female. According to the ASE/EACVI guidelines, LVDD was indeterminate in 5.9% and present in 8.4% of participants, of whom 54.3% had an indeterminate LVDD grade. Age, hypertension, diabetes mellitus, and cardiovascular disease were strongly associated with LVDD. Using the PBHS diastolic score, scores ≥3 and ≥4 were observed in 14.1% and 5.8% of the population, respectively. ASE/EACVI grades 2-3 corresponded with scores ≥4, whereas indeterminate categories corresponded with scores of 1-5. After multivariable adjustment, both LVDD grading systems predicted MACE (ASE/EACVI grade indeterminate and grades 2-3: aHRs 2.49 [95%CI: 1.39-4.46] and 5.7 [95%CI: 3.01-10.7], respectively; PBHS scores 4 and 5-6: aHRs 2.3 [95%CI: 1.25-4.33] and 5.3 [95%CI: 2.44-11.56], respectively).

**Conclusions:** This study validated the prognostic value of the ASE/EACVI diastolic function guidelines in the community. Assessing LVDD using a diastolic score free of indeterminate classifications may be worth exploring in future studies.

**ClinicalTrials.gov Identifier:** NCT03154346

## INTRODUCTION

Left ventricular diastolic dysfunction (LVDD) plays a central role in the assessment of patients with heart failure (HF), and dysfunction may manifest years to decades before HF symptoms appear.^1^ Community-based studies have previously shown that the presence of LVDD is a predictor of future cardiovascular outcomes.^2,3^ For example, in the Olmsted county study, Redfield et al. demonstrated that LVDD, often not accompanied by recognized HF, is associated with a marked increase in all-cause mortality.^2^ Additionally, in a Flemish community-based cohort, Kuznetsova et al. found that epidemiologically defined LVDD was an independent predictor of cardiovascular events after adjustment for risk factors.^3^

The mechanisms underlying LVDD may include impaired ventricular relaxation, increased chamber stiffness, and/or elevated ventricular filling pressures.^4^ These changes can be measured using echocardiographic parameters, including mitral inflow and tissue Doppler imaging (TDI) velocities, left atrial volume index (LAVI), and the presence of an elevated tricuspid regurgitation velocity (TRV).^5,6^ The American Society of Echocardiography (ASE) and European Association of Cardiovascular Imaging (EACVI) have developed diagnostic criteria for LVDD centered around four parameters: early mitral TDI velocity (e’), the ratio of early diastolic inflow velocity to e’ (E/e’), LAVI, and peak TRV.^6^ In patients with normal left ventricular ejection fraction (LVEF), LVDD is classified based on a simple majority, where LVDD is present if greater than 50% of criteria are met, absent if less than 50% of criteria are met, and indeterminate if exactly 50% of criteria are met. A second algorithm grades LVDD based on the ratio of mitral inflow velocities (E/A) and the presence of a majority of E/e’, LAVI, and TRV abnormalities, which is applicable to patients with reduced LVEF and those with LVDD and normal LVEF.^6^

While current ASE/EACVI criteria have demonstrated to be valuable in HF patients and hospitalized cohorts, very few studies have validated their prognostic utility in the general community.^3^ One limitation of the current algorithm is that a significant number of patients may remain unclassified or indeterminate, proportions of which have ranged from 20%-60% in different cohorts,^7–9^ leaving the diagnosis unclear. Moreover, in the community-based setting, TRV signal may not be available (ie, because of the lack of significant TR), limiting diagnostic yield.^10^ When absent, the simple majority system will implicitly assume TRV presence as abnormal, which could misclassify individuals. To get insights into these constraints and to quantify diastolic abnormalities, one may consider a scoring system based on minor and major criteria focusing on higher yield signals (eg, e’, E/e’, and LAVI) **(Central Illustration-A)**. As a precedent, the Heart Failure Association (HFA) of the European Society of Cardiology has previously employed minor and major criteria for echocardiography parameters within the HFA-PEFF score to diagnose heart failure with preserved ejection fraction (HFpEF).^11^

We leveraged data from the Project Baseline Health Study (PBHS), a longitudinal multicenter cohort study.^12^ In this sub-study, we first aimed to validate the prognostic value of LVDD assessed by the ASE/EACVI criteria in a community population. Second, we sought to explore how a PBHS diastolic score, incorporating minor and major criteria, would perform in quantifying the diastolic abnormalities present and predicting the risk of cardiovascular outcomes. We hypothesized that this would allow us to characterize and risk-stratify the challenging LVDD indeterminate categories (class and grade) of the guidelines.

## METHODS

### Study Population

The design of the PBHS study has been previously detailed.^12,13^ Briefly, the PBHS study enrolled 2,502 participants from an online registry of diverse adults. The study was conducted across multiple sites: Stanford University, Duke University, and the California Health and Longevity Institute, with enrolling sites in Palo Alto, California; Durham and Kannapolis, North Carolina; and Los Angeles, California. The study was approved by the Stanford University and Duke University Institutional Review Boards, and participants provided written informed consent before participation.

The inclusion and exclusion criteria for the analytic population in this PBHS sub-study are described in **Figure 1**. Considering the baseline visit, we excluded individuals without echocardiography as well as those with moderate or severe mitral valve disease, atrial fibrillation or flutter at the time of echocardiography, and a paced atrial or ventricular rhythm. Additionally, individuals who did not have simultaneously available e’, E/e’, and LAVI measurements at enrollment were excluded. The overall study cohort included 1,952 participants. Subsequently, we defined a subset of these individuals as a reference group to derive the thresholds considered for the scoring system **(Figure 1)**. The reference group consisted of individuals who had: (1) no current or prior history of cardiovascular, pulmonary, kidney, or autoimmune diseases or active cancers; (2) no major risk factors for atherosclerotic cardiovascular diseases, including hypertension, diabetes mellitus type II, obesity (body mass index [BMI] ≥30 kg/m^2^), dyslipidemia (low-density lipoprotein cholesterol ≥160 mg/dL) or active smoking; and (3) a coronary artery calcium (CAC) score <100 Agatson.

**Figure 1.**
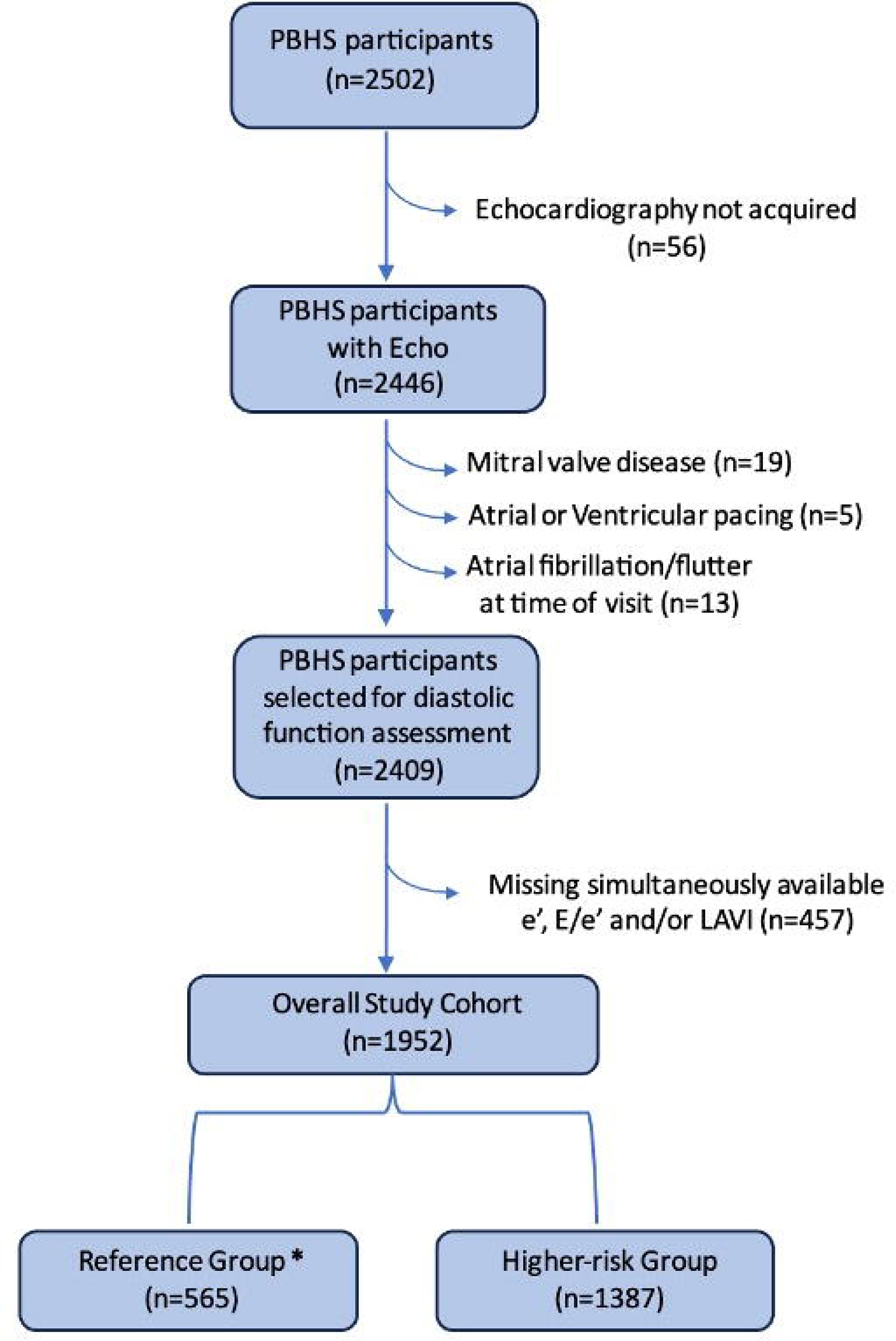
Flow chart of the Project Baseline Health Sub-Study population. The reference group excluded participants with established disease (cardiovascular, pulmonary, kidney, or autoimmune disease, or active cancers), major cardiovascular risk factors, and a coronary artery calcium score >100 Agatston.

### Echocardiographic Assessment

Echocardiograms were performed at each site, with the Duke Imaging Core Laboratory providing real-time quality control. Following a standardized protocol, echocardiography included M-mode, 2-dimensional, pulsed-wave Doppler and TDI imaging. Chamber quantification and assessment of diastolic function was performed by Imaging Core Laboratory personnel, blinded to patients’ clinical data, using the Digisonics software platform.

Measured echocardiographic parameters included left ventricular (LV) mass using the linear method, LV linear dimensions, biplane end-diastolic and end-systolic volumes, wall thickness, relative wall thickness, septal and lateral e’, early and late diastolic peak velocities of mitral inflow ratio (E/A), average E/e’, left atrial volumes (LAV), peak TRV, estimated right ventricular systolic pressure, and aortic root diameters. LAV and LV volumes were measured using the biplane Simpson method and scaled to body surface area. Intraclass correlation coefficients reported by the Core Laboratory reproducibility assessments were 0.94 for LV end-diastolic volume, 0.99 for LV mass, 0.96 for LV ejection fraction, and 0.94 for LAV.^13^

### Diastolic Dysfunction Assessment and Minor and Major Criteria of LVDD

We evaluated LVDD using the ASE/EACVI classification, which consists of a 2-step approach, yielding a diastolic dysfunction classification (present, absent, or indeterminate ∼*Algorithm A*) and a subsequent grading of the LVDD (grade 1, 2, 3, or grade indeterminate ∼*Algorithm B*). In the absence of myocardial disease or LVEF <50%, the following four criteria were evaluated: e’ velocity (septal e’ <7 cm/s, lateral e’ <10 cm/s), average E/e’ >14, peak TRV >2.8 cm/s, or LAVI >34 mL/m^2^. LVDD was present when >50% of the criteria were met; indeterminate, if exactly 50% of the criteria were met; and absent, if <50% of the criteria were met. In the presence of LVDD, an LVEF <50%, or myocardial disease, dysfunction was further graded according to the E/A ratio, E/e’, peak TRV, and LAVI, following the ASE/EACVI *Algorithm B.*^6^

Alternatively, we explored the performance of an empiric PBHS diastolic score. The reference values for the major and minor criteria of the score were guided by percentile-based cutoffs of the parameters e’, E/e’, and LAVI in the PBHS reference group. For major criteria thresholds, we used values ≤ 2.5^th^ percentile cut-off for septal and lateral e’, and ≥ 97.5^th^ percentile cut-off for E/e’ and LAVI. For minor criteria thresholds, we used values outside the 15^th^-85^th^ percentile ranges. Thresholds were rounded to the closest unit while ensuring consistency with previous recommendations, if applicable. Peak TRV was not included in the scoring system due to the frequent unavailability of this measurement. Additionally, TRV may lack specificity for diastolic dysfunction as elevated levels may also reflect pre-capillary pulmonary hypertension.^14^

### Outcome Assessment

Clinical events were reported by participants or confirmed by study coordinators using pre-specified visit case report forms. Major adverse cardiovascular events (MACE) were defined as a first cardiovascular-related event, which included composites of cardiovascular-related mortality, atherosclerotic events, incident HF, thrombotic events, new-onset atrial fibrillation, hypertensive emergency, pacemaker implantation, and valve replacement procedures. The median follow-up period was 4.25 years, and the time-to-first-event was considered for analysis. We excluded participants with a reported mortality from unknown cause if they did not have any preceding cardiovascular event (competing events).

### Statistical Analysis

The online Verily Terra platform (https://terra.bio/) was used to access the data, which were analyzed using the Python 3.10 environment. Baseline characteristics were presented for the overall cohort population and reference group. Continuous variables were summarized as means ± standard deviation and categorical variables as frequencies and percentages. To analyze the associations between LVDD and cardiovascular risk factors, a backward stepwise logistic regression analysis was performed. Odds ratios and 95% confidence intervals (CIs) were calculated and displayed as forest plots using a log_2_ scale. To evaluate the association of LVDD with incident MACE, the Kaplan–Meier method was employed to estimate event-free probabilities and graphically depict the time-to-event data for the duration of the study follow-up. This was followed by a univariable Cox regression screen **(Supplemental Table 1)** and a multivariable Cox regression analysis to estimate the proportional adjusted hazard ratios (aHRs) and 95% CIs for the risk of MACE according to the guidelines LVDD grading and PBHS diastolic scores. The following covariables were used in all the multivariable-adjusted models: age, sex, BMI, systolic blood pressure, active smoking status, serum high-density lipoprotein (HDL) cholesterol, non-HDL cholesterol, estimated glomerular filtration rate (eGFR), lipid-lowering medication use, hypertensive medication use, history of diabetes mellitus, history of cardiovascular disease (CVD), LVEF <50%, and LV mass index. For Cox regression (lifelines 0.27.8), using grid search optimization, the values 0.1, 0.05, and 0.01 were tested for the penalizer and L1 ratio hyperparameters, aiming at maximizing the average evaluation of the out-of-sample log-likelihood. Scaled Schoenfeld residuals were used to test proportional hazard assumptions.

To assess the clinical utility of including diastolic function evaluation in cardiovascular risk prediction, we compared a base clinical model composed of the same cardiovascular risk factors in the multivariable Cox analysis with an incremental model that incorporated the same variables plus LVDD grade/score. The comparison was conducted using likelihood-ratio chi-squared tests, c-statistics, and adjusted Brier scores.^15^

## RESULTS

### Study Sample Characteristics

**Table 1** presents the clinical and echocardiographic characteristics of the overall study cohort and the healthy reference group. The mean age of the participants (n=1,952) was 50±16 years; 56.3% were female; and 35.6% had hypertension, of whom 72.6% were on antihypertensive treatment. Only 31 (1.6%) individuals had an LVEF <50%. Compared with the overall study cohort, the reference sample was younger (41±14 vs. 50±16 years, p<0.001); however, both groups had similar racial and ethnic representation.

**Table 1.**
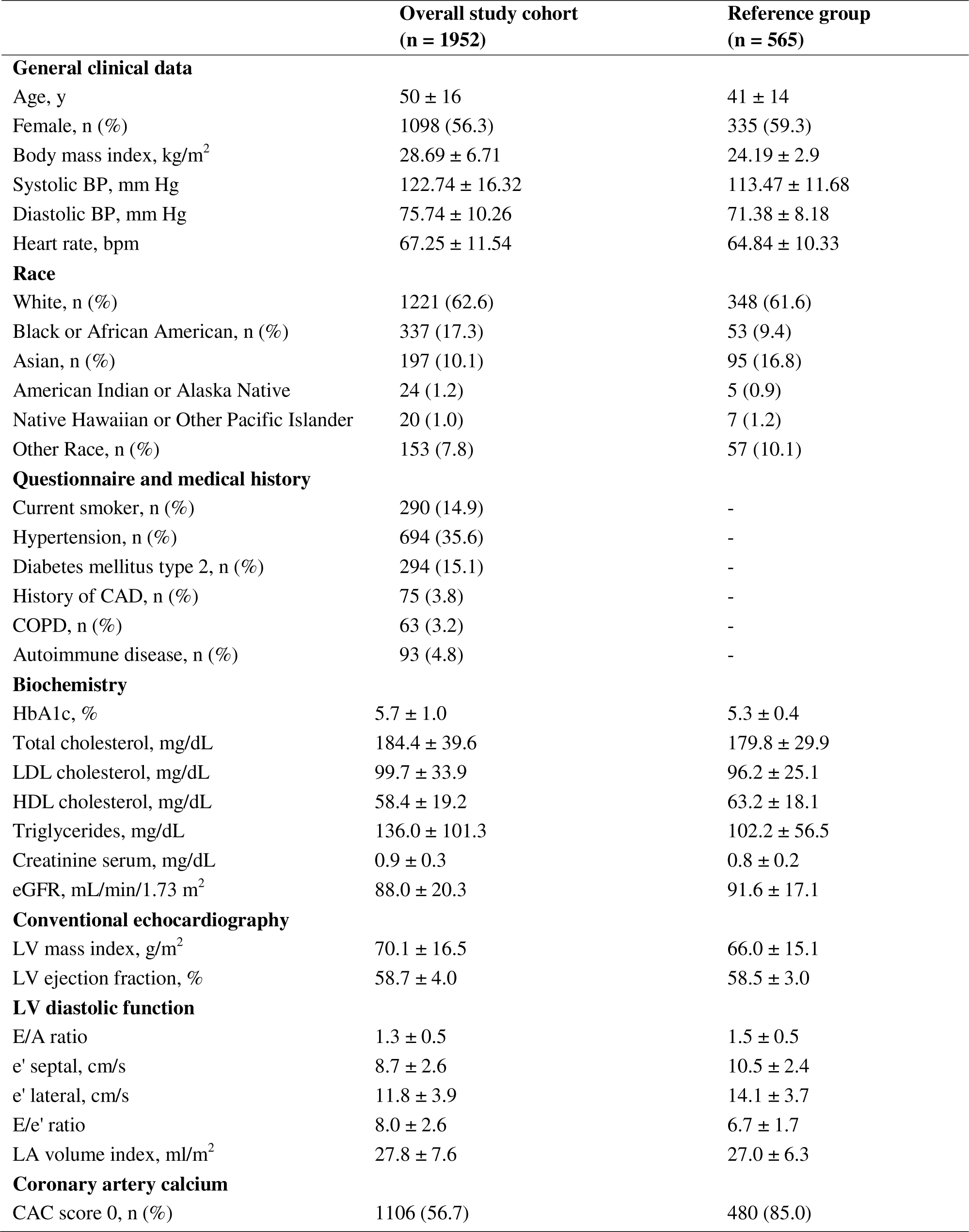
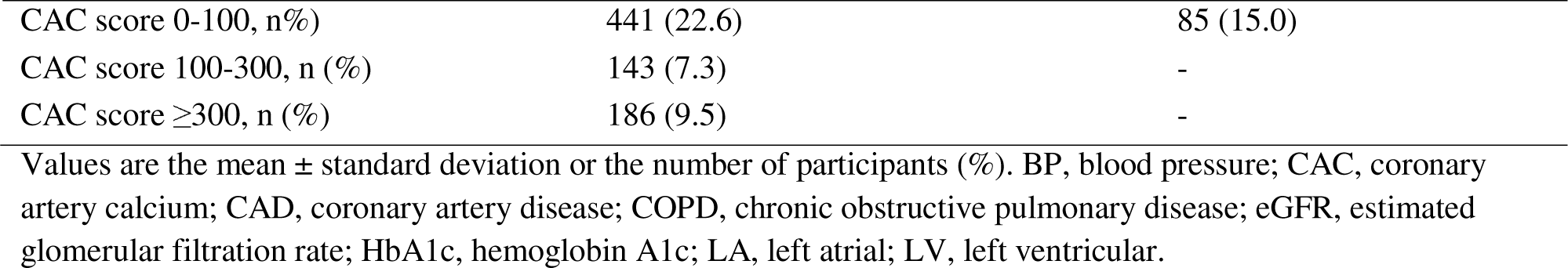
General characteristics of the study population.

|  | **Overall study cohort**<br>**(n = 1952)** | **Reference group**<br>**(n = 565)** |
| --- | --- | --- |
| **General clinical data** |  |  |
| Age, y | 50 ± 16 | 41 ± 14 |
| Female, n (%) | 1098 (56.3) | 335 (59.3) |
| Body mass index, kg/m<sup>2</sup> | 28.69 ± 6.71 | 24.19 ± 2.9 |
| Systolic BP, mm Hg | 122.74 ± 16.32 | 113.47 ± 11.68 |
| Diastolic BP, mm Hg | 75.74 ± 10.26 | 71.38 ± 8.18 |
| Heart rate, bpm | 67.25 ± 11.54 | 64.84 ± 10.33 |
| **Race** |  |  |
| White, n (%) | 1221 (62.6) | 348 (61.6) |
| Black or African American, n (%) | 337 (17.3) | 53 (9.4) |
| Asian, n (%) | 197 (10.1) | 95 (16.8) |
| American Indian or Alaska Native | 24 (1.2) | 5 (0.9) |
| Native Hawaiian or Other Pacific Islander | 20 (1.0) | 7 (1.2) |
| Other Race, n (%) | 153 (7.8) | 57 (10.1) |
| **Questionnaire and medical history** |  |  |
| Current smoker, n (%) | 290 (14.9) | - |
| Hypertension, n (%) | 694 (35.6) | - |
| Diabetes mellitus type 2, n (%) | 294 (15.1) | - |
| History of CAD, n (%) | 75 (3.8) | - |
| COPD, n (%) | 63 (3.2) | - |
| Autoimmune disease, n (%) | 93 (4.8) | - |
| **Biochemistry** |  |  |
| HbA1c, % | 5.7 ± 1.0 | 5.3 ± 0.4 |
| Total cholesterol, mg/dL | 184.4 ± 39.6 | 179.8 ± 29.9 |
| LDL cholesterol, mg/dL | 99.7 ± 33.9 | 96.2 ± 25.1 |
| HDL cholesterol, mg/dL | 58.4 ± 19.2 | 63.2 ± 18.1 |
| Triglycerides, mg/dL | 136.0 ± 101.3 | 102.2 ± 56.5 |
| Creatinine serum, mg/dL | 0.9 ± 0.3 | 0.8 ± 0.2 |
| eGFR, mL/min/1.73 m<sup>2</sup> | 88.0 ± 20.3 | 91.6 ± 17.1 |
| **Conventional echocardiography** |  |  |
| LV mass index, g/m<sup>2</sup> | 70.1 ± 16.5 | 66.0 ± 15.1 |
| LV ejection fraction, % | 58.7 ± 4.0 | 58.5 ± 3.0 |
| **LV diastolic function** |  |  |
| E/A ratio | 1.3 ± 0.5 | 1.5 ± 0.5 |
| e' septal, cm/s | 8.7 ± 2.6 | 10.5 ± 2.4 |
| e' lateral, cm/s | 11.8 ± 3.9 | 14.1 ± 3.7 |
| E/e' ratio | 8.0 ± 2.6 | 6.7 ± 1.7 |
| LA volume index, mL/m<sup>2</sup> | 27.8 ± 7.6 | 27.0 ± 6.3 |
| **Coronary artery calcium** |  |  |
| CAC score 0, n (%) | 1106 (56.7) | 480 (85.0) |
| CAC score 0-100, n(%) | 441 (22.6) | 85 (15.0) |
| CAC score 100-300, n (%) | 143 (7.3) | - |
| CAC score $\geq 300$, n (%) | 186 (9.5) | - |

Values are the mean ± standard deviation or the number of participants (%). BP, blood pressure; CAC, coronary artery calcium; CAD, coronary artery disease; COPD, chronic obstructive pulmonary disease; eGFR, estimated glomerular filtration rate; HbA1c, hemoglobin A1c; LA, left atrial; LV, left ventricular.

### LVDD According to the ASE/EACVI Recommendations

#### Prevalence of LVDD

In the overall cohort, LVDD was present in 164 participants (8.4%) while indeterminate classification of LVDD was observed in 115 participants (5.9%) according to the ASE/EACVI criteria. No LVDD was observed in the remaining participants. Among the participants with LVDD, grade 1 was identified in 34 individuals (20.7%), grade 2 in 36 individuals (21.9%), and grade 3 in 5 individuals (3.1%). However, an indeterminate grade was identified in the majority of participants with LVDD (54.3%) (**Figure 2)**. In the reference cohort, most participants (96.6%) did not have LVDD; 2.5% were classified as indeterminate for LVDD; and 0.9% met the criteria for LVDD but had an indeterminate grade **(Supplemental Figure 1)**. Of note, peak TRV signal was only present in 47% of the overall cohort.

**Figure 2.**
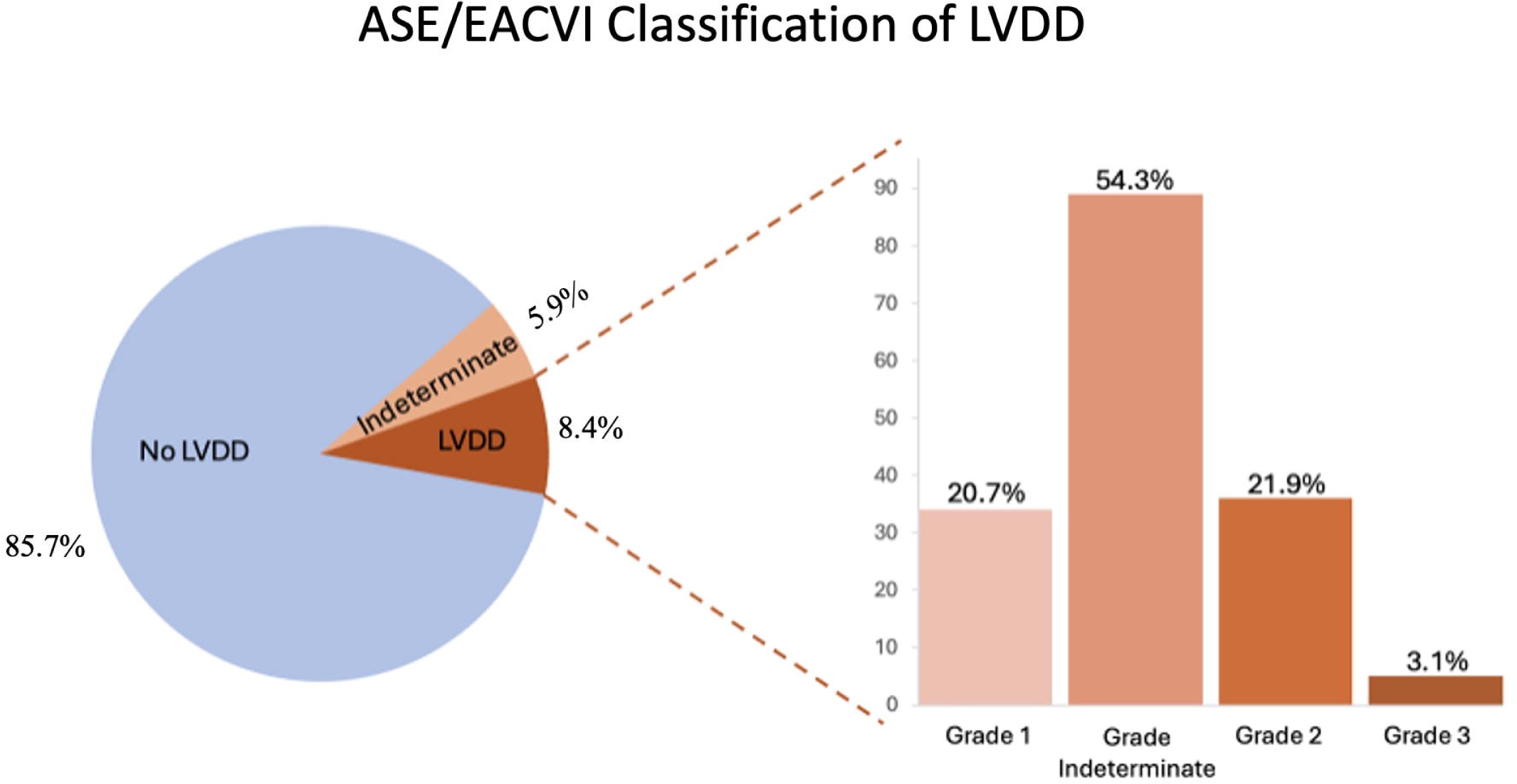
Distribution of LVDD categories according to the ASE/EACVI classification in the overall Project Baseline Health Study (PBHS) cohort.

#### Clinical Correlates of LVDD

LVDD, diagnosed by the ASE/EACVI criteria, was associated with age, pulse pressure, non-HDL cholesterol, diabetes mellitus, hypertension, and a history of CVD **(Figure 3)**. A history of CVD had the largest effect estimate, with a 5-fold higher odds of having LVDD (OR 5.13; 95% CI 3.41-7.70; p<0.001).

**Figure 3.**
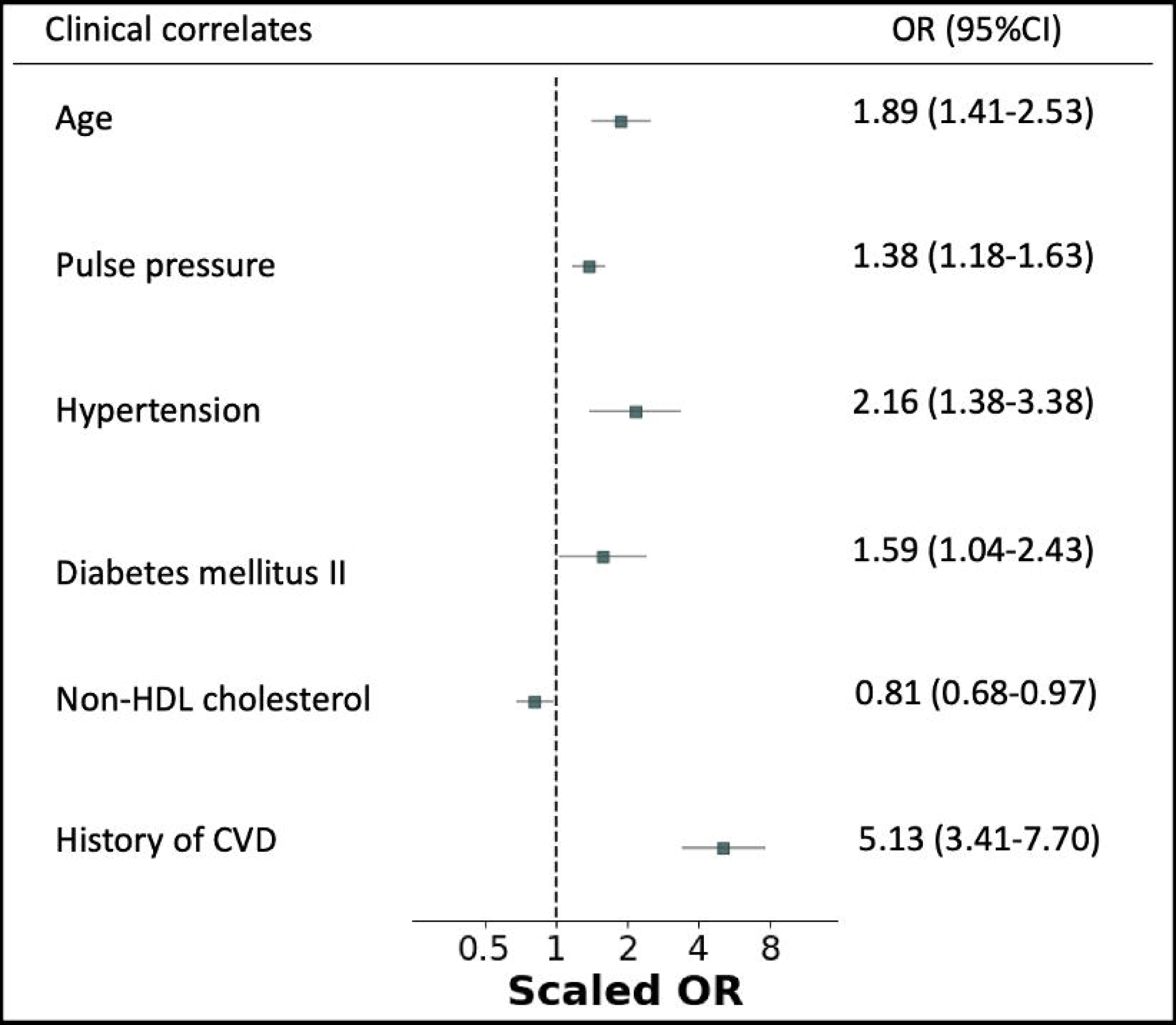
Association of clinical correlates with LVDD. Values represent the odds of having LVDD according to the ASE/EACVI criteria. Odds ratios of continuous values were scaled to half the 16th-84th percentile range, which is equal to 19 years for age, 11 mmHg for pulse pressure, 37 mg/dL for non-HDL cholesterol, and 19 mg/dL for HDL cholesterol.

#### LVDD and Major Adverse Cardiovascular Events

After a median follow-up of 4.25 years, 98 individuals experienced at least one MACE. Among them, 54.1% were atherosclerosis-related (n=53), and 26.6% were new-onset atrial fibrillation (n=26). The remaining MACE were composites of thrombotic events (8.2%), cardiovascular intervention (valve replacement or pacemaker implantation) (7.1%), incident HF (3.1%), hypertensive emergencies (1%), and cardiovascular-related mortality (1%). **Figure 4A** displays the Kaplan–Meier curves of cardiovascular event-free probabilities across the ASE/EACVI classification categories, suggesting an incremental worse prognosis with every LVDD grade increase. **Figure 4B** shows the univariable and multivariable aHRs for MACE based on the ASE/EACVI criteria. In multivariable models, individuals with an ASE/EACVI grade of indeterminate (HR 2.49; 95% CI 1.39-4.46; p=0.021) or grade 2 or 3 (HR 5.67; 95% CI 3.01-10.67; p<0.001) had a significantly higher risk of a MACE compared with individuals with no LVDD or an indeterminate LVDD class **(Figure 4B, Supplemental Table 2)**.

**Figure 4.**
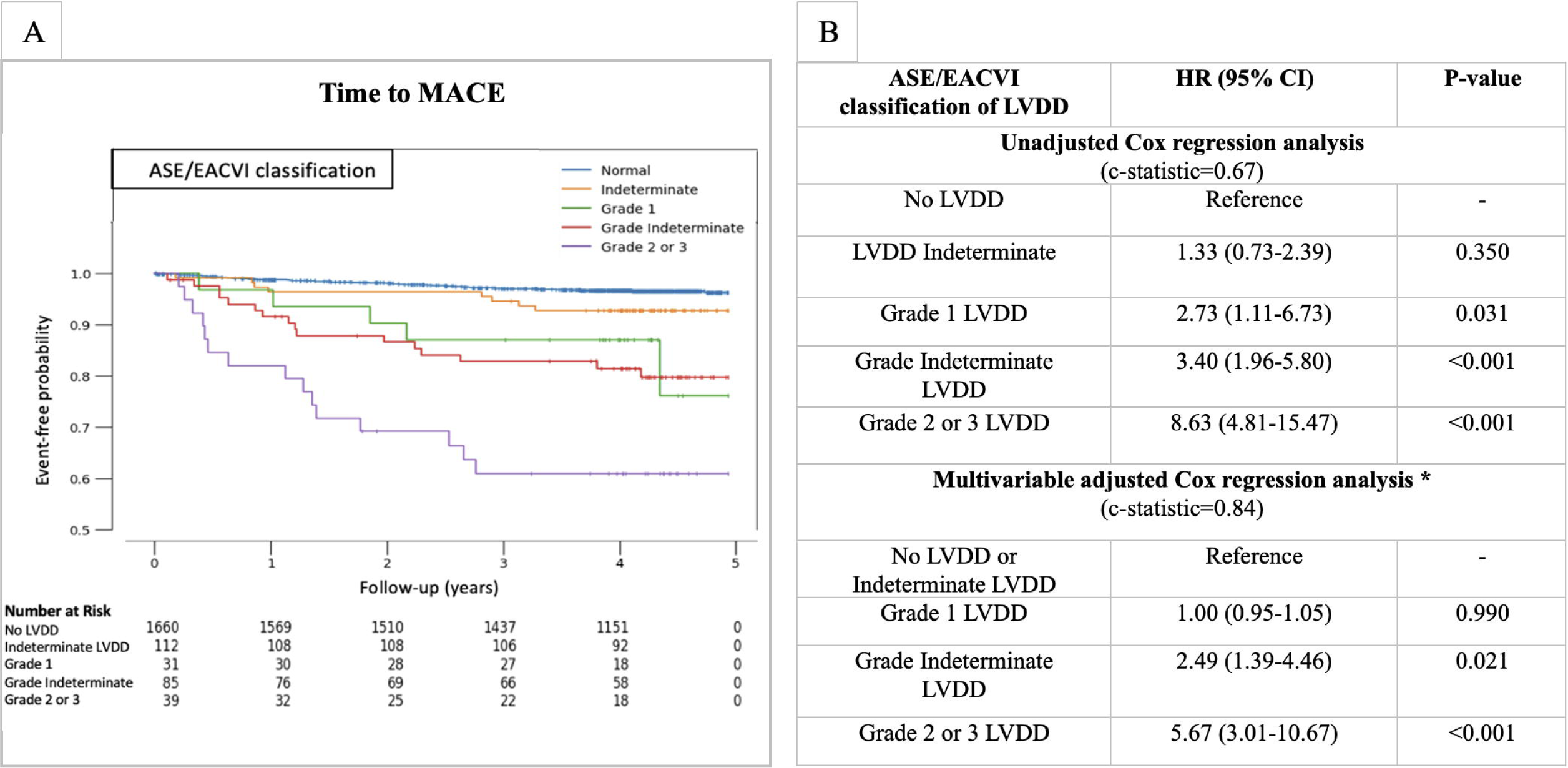
Associations of LVDD categories with major adverse cardiac events (MACE) according to the ASE/EACVI classification. (A) Kaplan–Meier curves of event-free probabilities. Grades 2-3 were combined due to the limited number of individuals in these categories. (B) Cox regression table for the risk of MACE per LVDD category. Hazard ratios (HRs) represent the relative hazard for each category compared with the reference group. The multivariable model was adjusted for age, sex, body mass index, active smoking, systolic blood pressure, high-density lipoprotein (HDL) cholesterol, non-HDL cholesterol, estimated glomerular filtration rate, diabetes mellitus history, cardiovascular disease history, hypertensive medication use, lipid-lowering medication use, left ventricular hypertrophy, and left ventricular ejection fraction <50%.

To evaluate the potential clinical significance of incorporating diastolic function assessment into cardiovascular risk prediction, we tested the added value that an incremental model, including LVDD, provides over a base model composed solely of the established cardiovascular risk factors adjusted for in the Cox analysis. The likelihood-ratio chi-squared test was significant between both models when adding the ASE/EACVI criteria (p<0.001). Moreover, the adjusted Brier score, reflecting predictive accuracy, increased from 12% for the base model to 16.6% when adding LVDD by the ASE/EACVI criteria (**Figure 5A)**. Scatter plots displayed in **Figure 5B** indicate that the incremental models’ effect on predicted risk is more pronounced in higher age groups compared with the base model.

**Figure 5.**
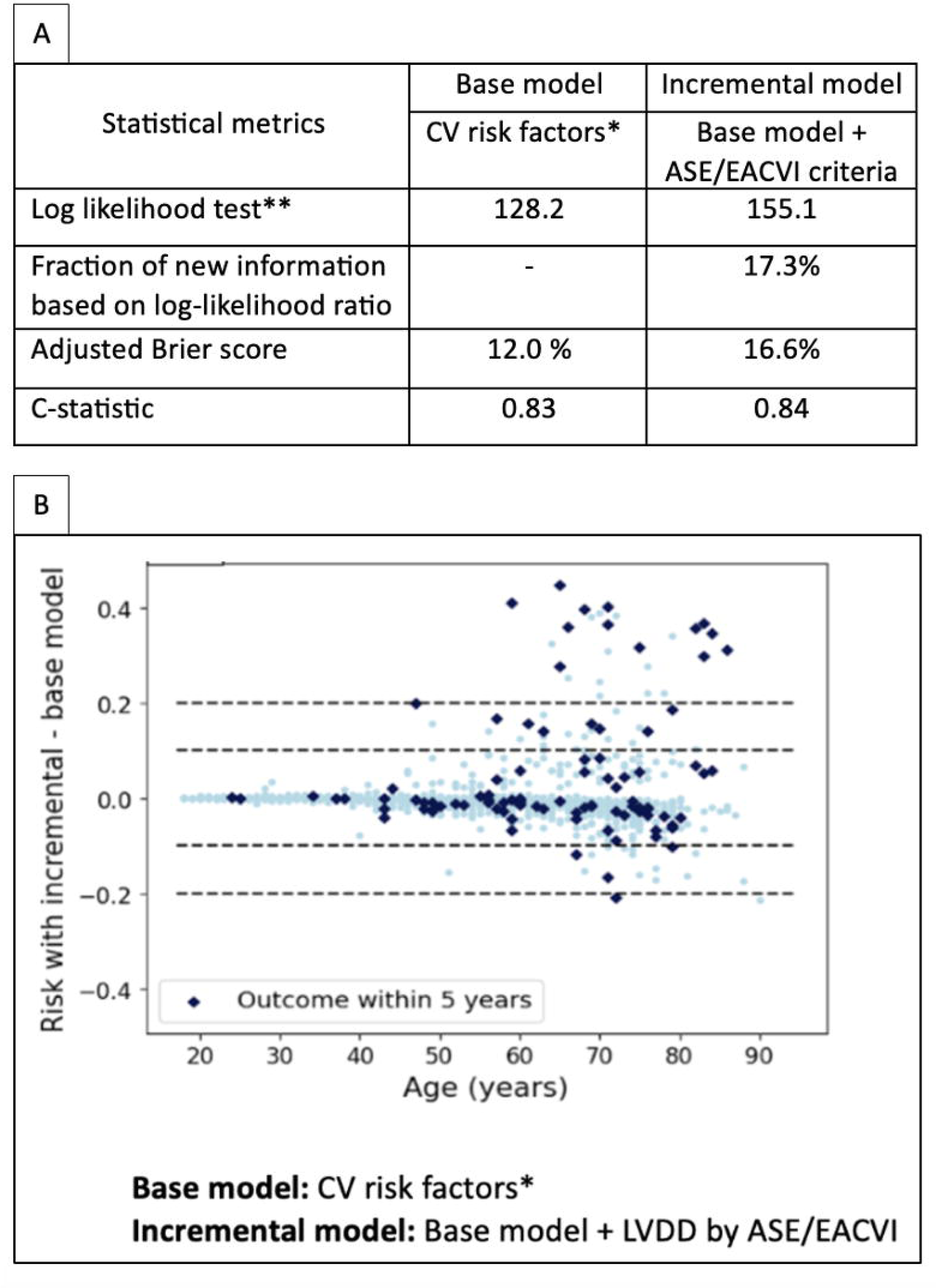
The value of incremental risk models considering LV diastolic function assessments in predicting MACE. (A) Statistical performance metrics of the base model supplemented with the ASE/EACVI criteria. (B) Scatter plot of the risk difference between the base model and the incremental model with the ASE/EACVI classification of left ventricular diastolic dysfunction (LVDD). Base model cardiovascular risk factors include age, sex, body mass index, systolic blood pressure, active smoking status, serum HDL cholesterol, non-HDL cholesterol, estimated glomerular filtration rate, lipid-lowering medication use, hypertensive medication use, history of diabetes mellitus, history of CVD, LVEF <50%, and LV mass index. P-value <0.001 for the difference in log likelihood between base and incremental model.

### The PBHS Diastolic Score

#### Score Criteria and Distribution in the PBHS Cohort

To capture an ordinal classification of diastolic function, we developed a PBHS diastolic score comprising minor and major criteria for e’, E/e’, and LAVI. Criteria were guided by the percentiles in the reference cohort (**Supplemental Table 3)**. The resultant PBHS diastolic scoring system is represented in **Figure 6A**. Minor criteria were empirically attributed 1 point in the scoring system and included septal e’ velocities of 5.5-7 cm/s, lateral e’ velocities of 7-10 cm/s, an average E/e’ of 10-14, and a LAVI of 34-42 ml/m^2^. Major criteria were empirically attributed 2 points each and included septal e’ velocities <5.5 cm/s, lateral e’ velocities <7 cm/s, an average E/e’ >14, and a LAVI >42 ml/m^2^. Although TRV was not included in the PBHS diastolic score due to its infrequent availability and poor specificity to LVDD,^10,15^ we tested its distribution in the PBHS population. The ASE/EACVI reference limit of 2.8 m/s corresponded to >99th percentile in our reference group **(Supplemental Table 3)**.

**Figure 6.**
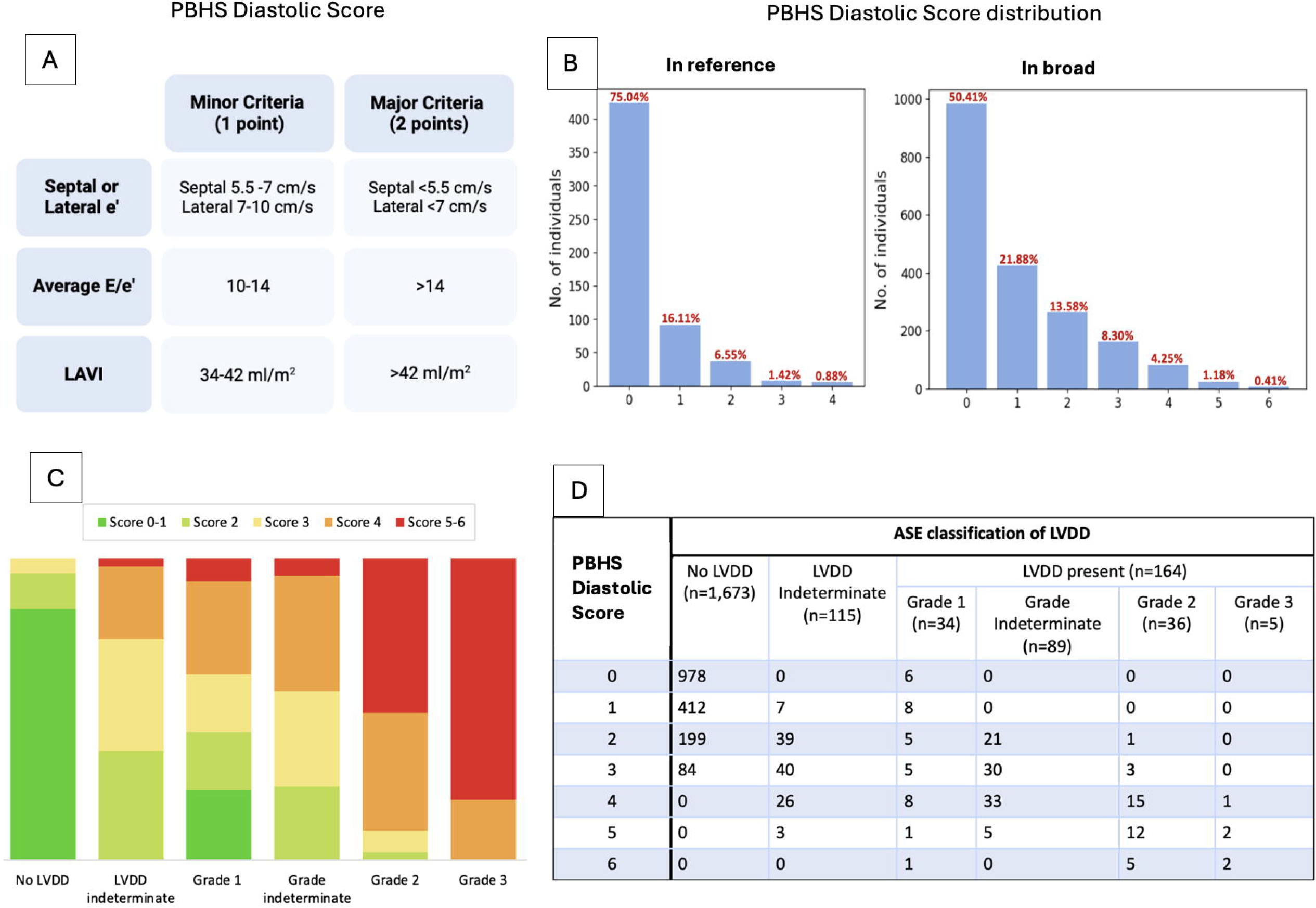
PBHS diastolic scoring system and overlap with ASE/EACVI classification of left ventricular diastolic dysfunction (LVDD). (A) PBHS diastolic score. (B) PBHS diastolic score distribution in the reference and broad cohorts. (C) Stacked bar-chart of the diastolic scores distribution across ASE/EACVI categories. (D) Cross-table showing the overlap of the ASE/EACVI classification with the PBHS diastolic scores.

In the overall cohort, PBHS diastolic scores of 0-1 were observed in most participants (72.3%), with 114 individuals (5.8%) having scores ≥4 **(Figure 6B)**. In the reference group, 552 participants (97.7%) had scores ≤2. When the diastolic scores were contrasted with the ASE/EACVI classification of LVDD **(Figures 3C, 3D)**, a clear concordance was noted. No participant with normal LV diastolic function according to the ASE/EACVI criteria had a PBHS diastolic score ≥4. Lower PBHS diastolic scores overlapped with no LVDD, whereas higher scores overlapped with higher grades (ASE/EACVI grades 2-3) of LVDD. In contrast, grade 1 and indeterminate class or grade of LVDD according to the ASE/EACVI criteria had scores ranging from low to high, without a clear clustering **(Figures 3C, 3D)**.

#### Clinical Correlates of the PBHS Diastolic Score

PBHS diastolic scores ≥4 (vs. <4) were associated with age, sex, pulse pressure, HDL cholesterol, non-HDL cholesterol, hypertension, and a history of CVD. Age had the most pronounced effect (OR 3.85; 95% CI 2.51-5.89; p<0.001) **(Supplemental Figure 2)**.

#### PBHS Diastolic Score and Major Adverse Cardiovascular Events

Kaplan–Meier curves of cardiovascular event-free probabilities across the PBHS diastolic scores **(Figure 7A)** suggest an incremental worse prognosis with every score point increase. In the multivariable Cox regression analysis, individuals with a PBHS diastolic score of 4 (HR 2.32; 95% CI 1.25-4.33; p=0.008) and scores of 5-6 (HR 5.32; 95% CI 2.44-11.56; p<0.001) were independently associated with a higher risk of MACE compared with individuals with scores of 2 or less **(Figure 7B, Supplemental Table 4)**.

**Figure 7.**
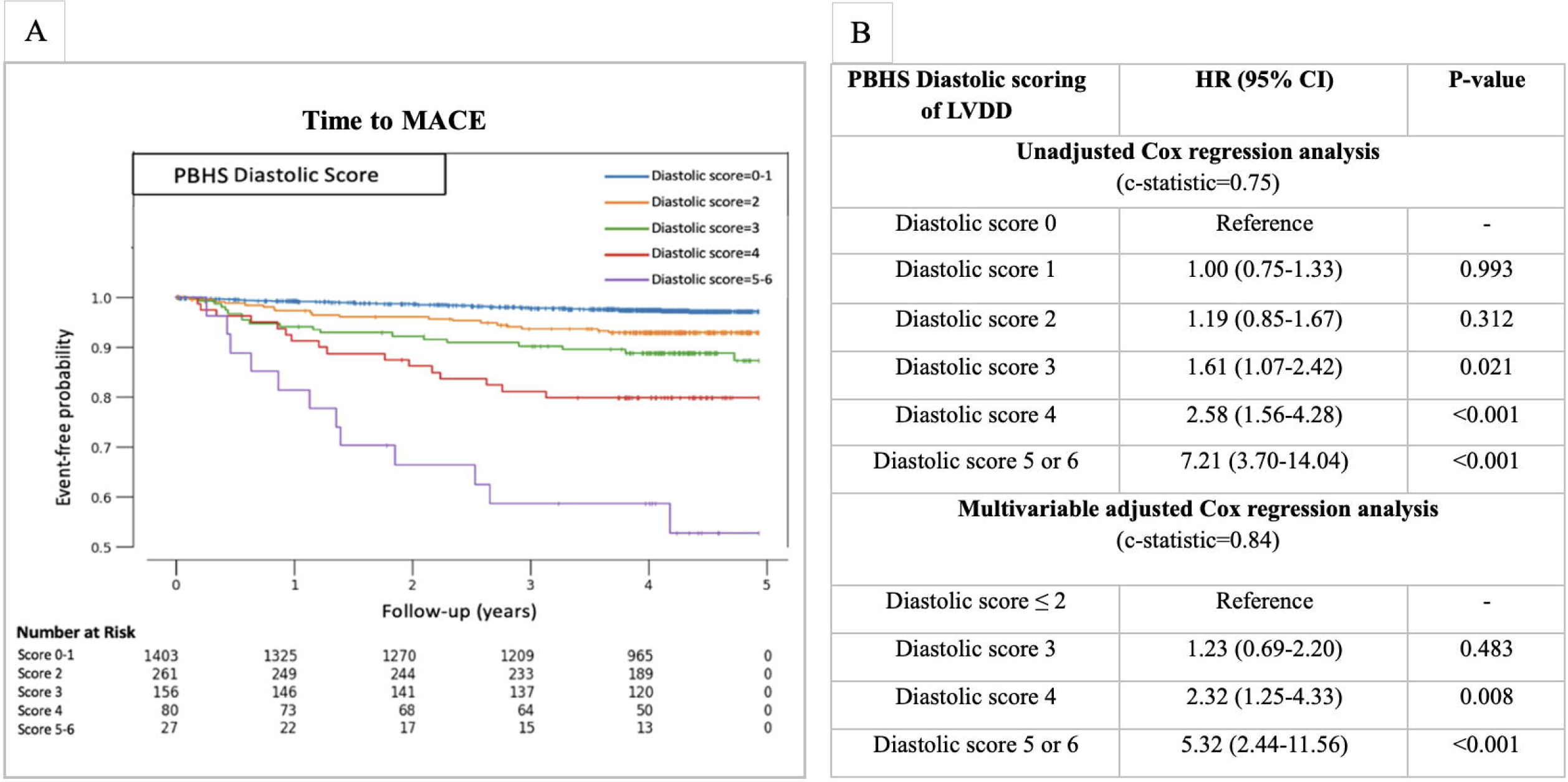
Associations of LVDD categories with major adverse cardiac events (MACE) according to the PBHS diastolic score. (A) Kaplan–Meier curves of event-free probabilities. Scores 0-1 were combined due to the scarcity of incident MACE. Scores 5-6 were combined due to the limited number of individuals in these categories. (B) Cox regression table for the risk of MACE per PBHS diastolic score. Hazard ratios (HRs) represent the relative hazard for each category compared with the reference group. The multivariable model was adjusted for age, sex, body mass index, active smoking, systolic blood pressure, high-density lipoprotein (HDL) cholesterol, non-HDL cholesterol, estimated glomerular filtration rate, diabetes mellitus history, cardiovascular disease history, hypertensive medication use, lipid-lowering medication use, left ventricular hypertrophy, and left ventricular ejection fraction <50%.

Similarly to LVDF assessed by the ASE/EACVI criteria, adding the PBHS diastolic score to a base model showed a significant improvement in the incremental model, particularly in higher age groups (**Supplemental Figure 3)**.

## DISCUSSION

Our study presents several main findings. First, in a community-based study with a multi-racial and multi-ethnic cohort, LVDD was relatively common and associated with age and cardiovascular risk factors. Second, advanced LVDD defined by the ASE/EACVI criteria was predictive of MACE in our cohort. Third, the proposed PBHS diastolic score allowed characterization of LVDD by quantifying diastolic abnormalities on an ordinal scale, thus providing insights into the risk stratification of patients classified in the indeterminate categories of the guidelines.

Reference limits play an important role in defining thresholds for diastolic parameters, and different cohorts were studied for this purpose.^3,16–18^ For LAVI, guidelines often recommend a threshold of 34 ml/m^2^ to define LA enlargement, which corresponded to the 90^th^ percentile of the NORRE cohort.^6,18^ In the PBHS cohort, we found a threshold of 42 mL/m^2^ for LAVI consistent with the cut-offs suggested by the World Alliance Societies of Echocardiography (WASE)^16^ (41 mL/m^2^) and FLEMENGHO^3^ (45 mL/m^2^) cohorts at their 97.5^th^ percentiles. For e’ and E/e’, choosing reference limits is more challenging in view of age-related changes, and guidelines have adopted a single threshold for simplicity. Our septal and lateral e’ thresholds were consistent with the younger age group in the WASE cohort, albeit higher values were found in older age groups. The CAC score phenotyping in the reference group excluded more severe subclinical cardiovascular disease than the WASE cohort, which could explain the higher e’velocities.^16^ Prior work from our group has demonstrated the association between LVDD and subclinical CAC.^13^ For the E/e’ ratio, a threshold may vary depending on the intended goal: either early detection of myocardial abnormality, high LV filling pressures, or associations with outcomes. The 2016 ASE/EACVI guidelines evolved to adopt a threshold of 14; however, various hemodynamic stress and epidemiologic studies support a lower threshold of 10.^2,5,19^ Diagnostic scores, such as the H2FPEF, used a cutoff of 9 to differentiate HFpEF from other dyspnea causes.^20^ Indeed, defining reference limits not only depends on the cohort, but also on the desired balance between the specificity and sensitivity of LVDD detection. This diagnostic balance offers clinicians options for selecting the optimal criteria to evaluate LVDD. For example, age- and sex-adjusted criteria may be preferred for the earlier detection of LVDD, particularly in young individuals.^3,17^ In contrast, by choosing single thresholds, the ASE/EACVI guidelines signify the adoption of age-weighted criteria, leading to a definite higher prevalence of LVDD in the elderly, with fewer false-positives in younger individuals.^6^ This study explored an LVDD scoring approach, derived from reference parameters, that combines highly specific and less specific thresholds as major and minor criteria, respectively.

In our study, the presence of LVDD defined using the ASE/EACVI criteria was not uncommon. As expected, we found that LVDD was associated with age and known cardiovascular risk factors. While LVDD has been thoroughly investigated in clinical studies of symptomatic HF patients, limited studies report the prognostic importance of LVDD in the community. In a recent study of individuals with risk factors, Potter et al. showed that in addition to clinical factors, global longitudinal strain and diastolic dysfunction (defined by E/e’>15, or E/e’>10 + LAVI >34mL/m^2^) were important to consider for better prediction of HF risk; as doing so reclassified 61% of individuals in the intermediate-risk group into the low-risk group.^21^ In the Flemish community cohort, Kuznetsova et al. found that LVDD defined by age- and sex-specific criteria correlated well with incident cardiovascular events, but not when graded using ASE/EACVI criteria.^3^ In the PBHS cohort, we found that both advanced ASE/EACVI grades (indeterminate, 2-3) and high diastolic scores (≥4 points) were independently associated with MACE after full adjustment for cardiovascular risk factors. Adding diastolic function to clinical factors for cardiovascular risk assessment had a modest, yet significant, improvement in cardiovascular outcome predictions, with the incremental value being more pronounced in older age groups. Future research is necessary to validate these findings and eventually inform targeted intervention strategies in patients that will benefit the most.

One caveat of the ASE/EACVI classification is the indeterminate categories that range from 20%-60% in the literature, with an unclear association with cardiovascular risk.^7–9^ Playford et al. reported that the indeterminate class and grade included up to 21.5% of individuals overall and 62.5% of individuals with reduced LVEF.^8^ Chao et al. also noted a significant prevalence of indeterminate categories (43.8%) among patients referred for echocardiography at the Mayo Clinic, which they reclassified into three distinct mortality risk clusters.^7^ In our cohort, indeterminate categories were common as well (6% of the total PBHS cohort had an indeterminate classification of LVDD and 54% of individuals with LVDD had an indeterminate grade). Moreover, their corresponding PBHS diastolic scores, reflecting the extent of diastolic abnormalities in the cohort, were heterogenous (1-5 points). This contrasts with LVDD grades 2 and 3 for which the corresponding scores were mostly high (≥4 points) and associated with a higher risk of MACE. Several studies trace indeterminate grading back to unmeasurable TRV.^22,23^ In PBHS participants, only 46% had TRV signals where peak TRV could be quantified with high confidence. This calls for considering a different approach in grading LVDD that could classify 100% of the individuals and provides a clear risk stratification. For example, the PBHS diastolic score, by adopting minor and major criteria and excluding TRV, allowed characterization of indeterminate categories by quantifying their diastolic abnormalities and their subsequent association with MACE, with good predictive accuracy. This approach is therefore worth considering in future studies, with the caveat that outcome studies should guide the weight attributed to the different parameters. If TRV were to be included, a smaller weight compared to the other specific diastolic parameters would be optimal for assessing LVDD.

### Limitations

Our analysis had several limitations. First, the PBHS diastolic score was empirically weighted with insights from epidemiology distribution, previous literature, and guidelines. Being cohort-dependent, our results benefit from further validation studies in larger population samples with varying comorbidities and a higher incidence of HF-related outcomes. Additionally, a follow-up period of more than 5 years may be needed to fully identify late-onset cardiovascular events from long-term progression of LVDD.

Despite these limitations, the PBHS cohort does exhibit several strengths, including a multicenter community cohort, a thorough core laboratory analysis, a study population enriched in metabolic risk factors, and diverse representation from multiple ethnicities and races.

## CONCLUSIONS

Our study validates the prognostic value of LVDD in a community setting and demonstrates that the PBHS diastolic score can quantify diastolic abnormalities and provide insights into risk stratification, especially for patients lacking TRV measurements or patients who fall into indeterminate LVDD categories.

## Author Contributions

GF, ES, NC, KMA, AFH, MAD, SHS, PSD, TK, MS. and FH conceptualized and designed the study, provided overall supervision, and critically revised the manuscript. GF, ES, and FH conducted the data analysis, interpreted the results, and drafted the initial manuscript. BC, MKC, SAS, SPB were involved in the acquisition of data and critically reviewed the manuscript. All authors have approved the final article and agree to be accountable for all aspects of the work.

## Data Availability Statement

The deidentified PBHS data corresponding to this study are available upon request for the purpose of examining its reproducibility. Requests are subject to approval by PBHS governance.

## Tweet

In this Project Baseline Health Study, researchers validated #LVDD guidelines in the community population and investigated the potential use of a novel diastolic score to characterize LVDD and gain insights on the diastolic abnormalities in patients that could not be classified (indeterminate) per the guidelines @StanfordCVI @DCRINews @Verily

## ABBREVIATIONS

ASE: American Society of Echocardiography
CAC: coronary artery calcium
CVD: cardiovascular disease
E: mitral inflow velocity
E/A: early and late diastolic peak velocities of mitral inflow
E/e’: mitral inflow to annular velocity ratio
EACVI: European Association of Cardiovascular Imaging
eGFR: estimated glomerular filtration rate
e’: early diastolic mitral annular velocity
FLEMENGHO: Flemish Study on Environment, Genes and Health Outcomes
HFA: Heart Failure Association
HF: heart failure
HFpEF: heart failure with preserved ejection fraction
HDL-C: high-density lipoprotein-C
LAV: left atrial volume
LAVI: left atrial volume index
LVDD: left ventricular diastolic dysfunction
LVEF: left ventricular ejection fraction
MACE: major adverse cardiovascular events
NORRE: Normal Reference Ranges for Echocardiography
PBHS: Project Baseline Health Study
TDI: tissue doppler imaging
TRV: tricuspid regurgitation velocity
WASE: World Alliance Societies of Echocardiography

## ACKNOWLEDGMENTS

The authors wish to thank Project Baseline Health Study participants and study sites.

---

**Central Illustration.**
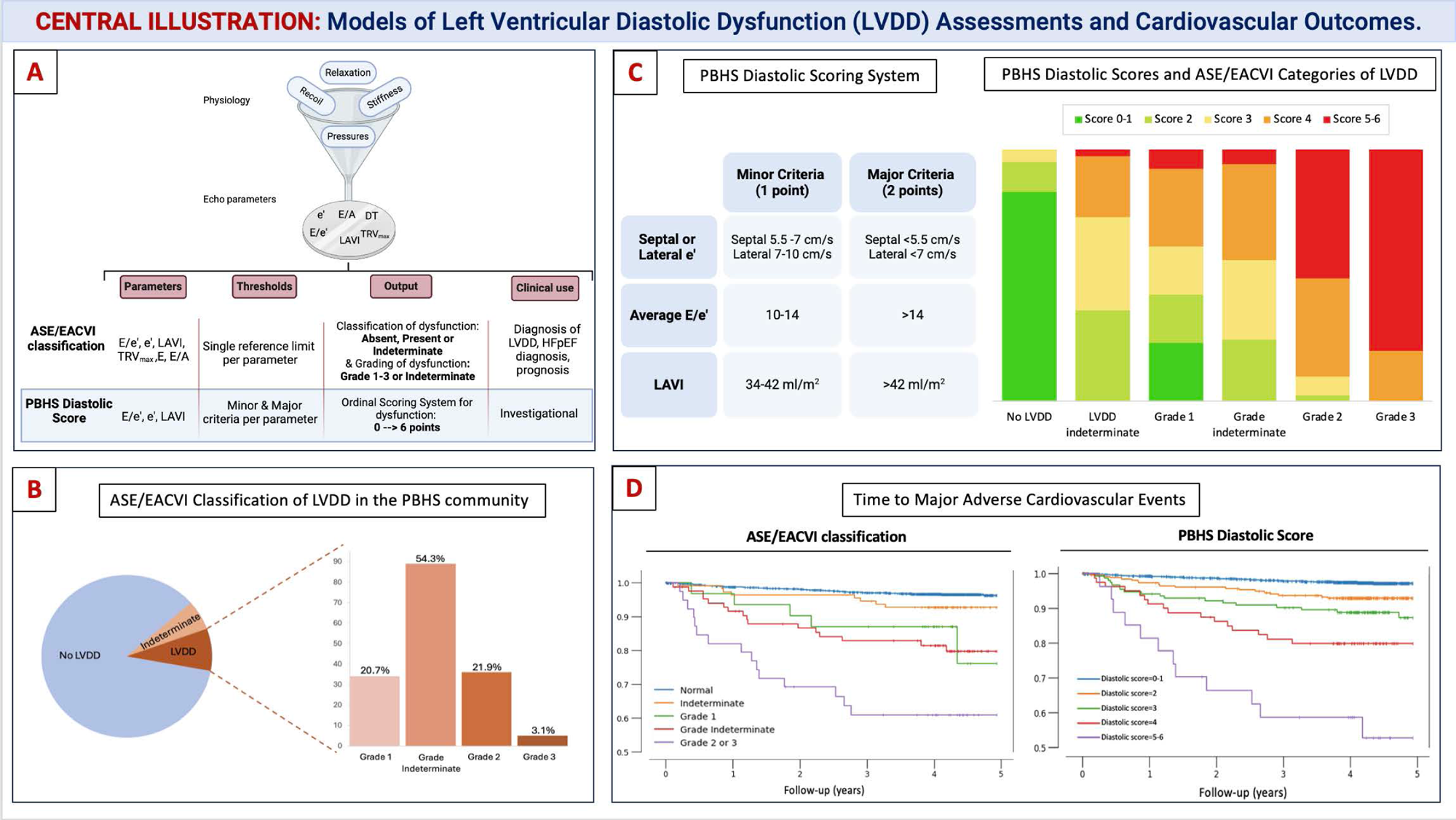
Models of Left Ventricular Diastolic Dysfunction (LVDD) Assessments and Cardiovascular Outcomes. (A) Concept figure: LVDD from physiology to clinical use. (B) Distribution of the American Society of Echocardiography (ASE) and European Association of Cardiovascular Imaging (EACVI) LVDD categories in the Project Baseline Health Study (PBHS). (C) PBHS diastolic scoring system and overlap with ASE/EACVI LVDD categories. (D) Kaplan–Meier of survival estimates according to the ASE/EACVI classification and PBHS diastolic score. DT, deceleration time; e’, early mitral tissue Doppler velocity; E/A, early diastolic filling velocity to atrial contraction filling velocity ratio; E/e’, the ratio of early diastolic inflow velocity to e’; HFA, Heart Failure Association; HFpEF, heart failure with preserved ejection fraction; GLS, global longitudinal strain; LAVI, left atrial volume index; LVMI, left ventricular mass index; RWT, relative wall thickness.

## REFERENCES

1. Heidenreich PA, Bozkurt B, Aguilar D, et al. 2022 AHA/ACC/HFSA Guideline for the Management of Heart Failure: A Report of the American College of Cardiology/American Heart Association Joint Committee on Clinical Practice Guidelines. Circulation. 2022;145(18). doi:10.1161/CIR.0000000000001063

2. Redfield MM, Jacobsen SJ, Burnett JC, Mahoney DW, Bailey KR, Rodeheffer RJ. Burden of Systolic and Diastolic Ventricular Dysfunction in the Community: Appreciating the Scope of the Heart Failure Epidemic. JAMA. 2003;289(2):194. doi:10.1001/jama.289.2.194

3. Kuznetsova T, Cauwenberghs N, Sabovčik F, Kobayashi Y, Haddad F. Evaluation of diastole by echocardiography for detecting early cardiac dysfunction: an outcome study. ESC Heart Fail. 2022;9(3):1775–1783. doi:10.1002/ehf2.13863

4. Zile MR, Brutsaert DL. New Concepts in Diastolic Dysfunction and Diastolic Heart Failure: Part II: Causal Mechanisms and Treatment. Circulation. 2002;105(12):1503–1508. doi:10.1161/hc1202.105290

5. Chetrit M, Cremer PC, Klein AL. Imaging of Diastolic Dysfunction in Community-Based Epidemiological Studies and Randomized Controlled Trials of HFpEF. JACC: Cardiovascular Imaging. 2020;13(1):310–326. doi:10.1016/j.jcmg.2019.10.022

6. Nagueh SF, Smiseth OA, Appleton CP, et al. Recommendations for the Evaluation of Left Ventricular Diastolic Function by Echocardiography: An Update from the American Society of Echocardiography and the European Association of Cardiovascular Imaging. Eur Heart J Cardiovasc Imaging. 2016;17(12):1321–1360. doi:10.1093/ehjci/jew082

7. Chao CJ, Kato N, Scott CG, et al. Unsupervised Machine Learning for Assessment of Left Ventricular Diastolic Function and Risk Stratification. Journal of the American Society of Echocardiography. 2022;35(12):1214–1225.e8. doi:10.1016/j.echo.2022.06.013

8. Playford D, Strange G, Celermajer DS, et al. Diastolic dysfunction and mortality in 436 360 men and women: the National Echo Database Australia (NEDA). Eur Heart J Cardiovasc Imaging. 2021;22(5):505–515. doi:10.1093/ehjci/jeaa253

9. Almeida JG, Fontes-Carvalho R, Sampaio F, et al. Impact of the 2016 ASE/EACVI recommendations on the prevalence of diastolic dysfunction in the general population. European Heart Journal - Cardiovascular Imaging. 2018;19(4):380–386. doi:10.1093/ehjci/jex252

10. O’Leary JM, Assad TR, Xu M, et al. Lack of a Tricuspid Regurgitation Doppler Signal and Pulmonary Hypertension by Invasive Measurement. JAHA. 2018;7(13):e009362. doi:10.1161/JAHA.118.009362

11. Pieske B, Tschöpe C, de Boer RA, et al. How to diagnose heart failure with preserved ejection fraction: the HFA-PEFF diagnostic algorithm: a consensus recommendation from the Heart Failure Association (HFA) of the European Society of Cardiology (ESC). Eur Heart J. 2019;40(40):3297–3317. doi:10.1093/eurheartj/ehz641

12. Arges K, Assimes T, Bajaj V, et al. The Project Baseline Health Study: a step towards a broader mission to map human health. npj Digit Med. 2020;3(1):84. doi:10.1038/s41746-020-0290-y

13. Haddad F, Cauwenberghs N, Daubert MA, et al. Association of left ventricular diastolic function with coronary artery calcium score: A Project Baseline Health Study. Journal of Cardiovascular Computed Tomography. 2022;16(6):498–508. doi:10.1016/j.jcct.2022.06.003

14. Lam CSP, Roger VL, Rodeheffer RJ, Borlaug BA, Enders FT, Redfield MM. Pulmonary Hypertension in Heart Failure With Preserved Ejection Fraction. Journal of the American College of Cardiology. 2009;53(13):1119–1126. doi:10.1016/j.jacc.2008.11.051

15. Harrell F. Statistically efficient ways to quantify added predictive value of new measurements. Published October 17, 2018. Accessed February 12, 2024.

16. Miyoshi T, Addetia K, Citro R, et al. Left Ventricular Diastolic Function in Healthy Adult Individuals: Results of the World Alliance Societies of Echocardiography Normal Values Study. Journal of the American Society of Echocardiography. 2020;33(10):1223–1233. doi:10.1016/j.echo.2020.06.008

17. Nayor M, Cooper LL, Enserro DM, et al. Left Ventricular Diastolic Dysfunction in the Community: Impact of Diagnostic Criteria on the Burden, Correlates, and Prognosis. JAHA. 2018;7(11):e008291. doi:10.1161/JAHA.117.008291

18. Kou S, Caballero L, Dulgheru R, et al. Echocardiographic reference ranges for normal cardiac chamber size: results from the NORRE study. European Heart Journal - Cardiovascular Imaging. 2014;15(6):680–690. doi:10.1093/ehjci/jet284

19. Nagueh SF, Middleton KJ, Kopelen HA, Zoghbi WA, Quiñones MA. Doppler Tissue Imaging: A Noninvasive Technique for Evaluation of Left Ventricular Relaxation and Estimation of Filling Pressures. Journal of the American College of Cardiology. 1997;30(6):1527–1533. doi:10.1016/S0735-1097(97)00344-6

20. Reddy YNV, Carter RE, Obokata M, Redfield MM, Borlaug BA. A Simple, Evidence-Based Approach to Help Guide Diagnosis of Heart Failure With Preserved Ejection Fraction. Circulation. 2018;138(9):861–870. doi:10.1161/CIRCULATIONAHA.118.034646

21. Potter E, Huynh Q, Haji K, et al. Use of Clinical and Echocardiographic Evaluation to Assess the Risk of Heart Failure. JACC: Heart Failure. 2024;12(2):275–286. doi:10.1016/j.jchf.2023.06.014

22. Balaney B, Medvedofsky D, Mediratta A, et al. Invasive Validation of the Echocardiographic Assessment of Left Ventricular Filling Pressures Using the 2016 Diastolic Guidelines: Head-to-Head Comparison with the 2009 Guidelines. Journal of the American Society of Echocardiography. 2018;31(1):79–88. doi:10.1016/j.echo.2017.09.002

23. Luke P, Eggett C, Spyridopoulos I, Irvine T. A comparative analysis of British and American Society of Echocardiography recommendations for the assessment of left ventricular diastolic function. Echo Res Pract. 2018;5(4):139–147. doi:10.1530/ERP-18-0024

