## Supplemental Material for "Prognostic Significance of Left Ventricular Diastolic Dysfunction in the Community: A Project Baseline Health Study"

| **Supplemental Table 1. Cox univariate hazard analysis of clinical variables and imaging-related factors and their association with major adverse cardiac events (MACE)** | **2-3** |
| --- | --- |
| **Supplemental Table 2. Full multivariable Cox regression analysis of the ASE/EACVI classification of left ventricular diastolic dysfunction (LVDD) in predicting MACE** | **4** |
| **Supplemental Table 3. Diastolic parameter values and corresponding percentiles in the Project Baseline Health Study (PBHS) reference group** | **5** |
| **Supplemental Table 4. Full multivariable Cox regression analysis of the PBHS diastolic score in predicting MACE** | **6** |
| **Supplemental Figure 1. (A) LVDD classification per the ASE/EACVI algorithm in the reference group. (B) Age- and sex-stratified distribution of individuals with LVDD in the reference group.** | **7** |
| **Supplemental Figure 2. Association of clinical correlates with LVDD using the PBHS diastolic score** | **8** |
| **Supplemental Figure 3. The value of incremental risk models considering LV diastolic function (LVDF) assessment by the PBHS diastolic score in predicting MACE** | **9** |

**Supplemental Table 1. Cox univariate hazard analysis of clinical variables and imaging-related factors** **and their association with major adverse cardiac events (MACE)**

| **Clinical variables** | **Hazard ratio** | **95% CI** | **P-value** |
| --- | --- | --- | --- |
| **Demographic and anthropometric** |  |  |  |
| Age | 2.49 | 1.99-3.17 | <0.001 |
| Male sex | 1.32 | 0.10-1.76 | 0.051 |
| BMI ≥ 30 | 1.09 | 0.81-1.47 | 0.562 |
| **Blood pressure** |  |  |  |
| Hypertension | 3.31 | 2.29-4.79 | <0.001 |
| Systolic blood pressure | 1.11 | 1.01-1.26 | 0.045 |
| **Metabolic** |  |  |  |
| Diabetes mellitus | 2.39 | 1.58-3.62 | <0.001 |
| HbA1C | 1.13 | 1.07-1.20 | <0.001 |
| Dyslipidemia | 2.41 | 1.66-3.49 | <0.001 |
| Non-HDL cholesterol | 0.96 | 0.80-1.12 | 0.560 |
| Triglycerides | 1.07 | 1.00-1.14 | 0.073 |
| HDL cholesterol | 0.71 | 0.57-0.88 | 0.001 |
| Ln (TG/HDL) | 1.33 | 1.12-1.59 | 0.001 |
| **Renal function** |  |  |  |
| Renal dysfunction (eGFR <60 mL/min/m^2^) | 1.88 | 0.92-3.83 | 0.085 |
| eGFR | 0.64 | 0.53-0.76 | <0.001 |
| Albumin/creatinine (urine) | 1.01 | 0.99-1.02 | 0.352 |
| **Lifestyle** |  |  |  |
| Current smoker | 1.90 | 1.22-2.95 | 0.005 |
| **CVD history** |  |  |  |
| Combined CVD | 3.48 | 2.40-5.03 | <0.001 |
| **Imaging factors** |  |  |  |
| LVEF <50% | 2.04 | 0.88-4.72 | 0.095 |
| LV hypertrophy | 2.47 | 1.51-4.05 | 0.003 |
| **PBHS diastolic score criteria** |  |  |  |
| e’ minor | 1.28 | 0.99-1.65 | 0.053 |
| e’ major | 2.63 | 1.85-3.76 | <0.001 |
| E/e’ minor | 1.41 | 1.04-1.92 | 0.029 |
| E/e’ major | 3.85 | 2.30-6.45 | <0.001 |
| LAVI minor | 1.74 | 1.09-2.78 | 0.021 |
| LAVI major | 2.56 | 1.51-4.32 | 0.001 |
| **Coronary artery calcium (CAC) score** |  |  |  |
| CAC 0 | 0.28 | 0.19 to 0.41 | <0.001 |
| CAC 1-100 | 1.0 | 0.996 to 1.00 | 0.999 |
| CAC 100-300 | 1.98 | 1.14 to 3.44 | 0.015 |
| CAC >300 | 3.69 | 2.52 to 5.40 | <0.001 |
| Ln (CAC+1) | 2.07 | 1.76 to 2.43 | <0.001 |

Hazard ratios of continuous variables were scaled to half the 16^th^-84^th^ percentile range.
BMI, body mass index; CI, confidence interval; CVD, cardiovascular disease; e', early diastolic mitral annular velocity; E/e', mitral inflow to annular velocity ratio; eGFR, estimated glomerular filtration rate; HDL, high-density lipoprotein; LAVI, left atrial volume index; LV, left ventricular; LVEF, left ventricular ejection fraction; TG, triglyceride.

**Supplemental Table 2. Full multivariable Cox regression analysis of ASE/EACVI classification of left ventricular diastolic dysfunction in predicting MACE**

|  | **Hazard ratio** | **95% CI** | **P-value** |
| --- | --- | --- | --- |
| ASE/EACVI Grade 1 | 1.00 | 0.95-1.05 | 1.000 |
| ASE/EACVI Grade Indeterminate | 2.49 | 1.39-4.46 | 0.002 |
| ASE/EACVI Grade 2 or 3 | 5.67 | 3.01-10.67 | <0.001 |
| Age at enrollment | 1.80 | 1.30-2.49 | 0.004 |
| Male sex | 1.29 | 0.86-1.92 | 0.209 |
| Body mass index | 0.91 | 0.75-1.11 | 0.355 |
| Systolic blood pressure | 1.03 | 0.86-1.23 | 0.748 |
| Active smoking | 2.35 | 1.46-3.79 | <0.001 |
| Estimated glomerular filtration rate | 0.96 | 0.77-1.19 | 0.711 |
| HDL cholesterol | 0.79 | 0.63-0.99 | 0.037 |
| Non-HDL cholesterol | 1.04 | 0.88-1.25 | 0.625 |
| Hypertensive medication | 1.19 | 0.75-1.88 | 0.457 |
| Lipid lowering medication | 1.14 | 0.73-1.77 | 0.568 |
| CVD history | 2.27 | 1.41-3.64 | <0.001 |
| Diabetes mellitus history | 1.60 | 1.01-2.54 | 0.046 |
| Ejection fraction <50% | 1.62 | 0.60-4.42 | 0.341 |
| Left ventricular hypertrophy | 1.93 | 1.10-3.38 | 0.019 |

Hazard ratios of continuous variables were scaled to half the 16^th^-84^th^ percentile range.

CI, confidence interval; CVD, cardiovascular disease; HDL, high-density lipoprotein.

**Supplemental Table 3. Diastolic parameter values and corresponding percentiles in the Project Baseline Health Study (PBHS) reference group**

| **Percentile** | **Septal e’** | **Lateral e’** | **Percentile** | **Average E/e’** | **LAVI** | **Peak TRV** |
| --- | --- | --- | --- | --- | --- | --- |
| 50 | 10.6 | 14.3 | >99 | 14 | 51.6 | 2.8 |
| 15 | 7.6 | 10.1 | 99 | 11.6 | 45.2 | 2.7 |
| 10 | 7 | 9.1 | 97.5 | 10.7 | 42 | 2.6 |
| 5 | 6.6 | 8 | 95 | 10 | 38 | 2.5 |
| 2.5 | 6 | 7.4 | 90 | 9.1 | 34 | 2.45 |
| 1 | 5.5 | 6.3 | 50 | 6.4 | 17.4 | 2 |

e’, early mitral tissue doppler velocity; E/e’, the ratio of early diastolic inflow velocity to e’; LAVI, left atrial volume index; TRV, tricuspid regurgitation velocity.

**Supplemental Table 4. Full multivariable Cox regression analysis of the PBHS diastolic score in predicting MACE**

|  | **Hazard ratio** | **95% CI** | **P-value** |
| --- | --- | --- | --- |
| Diastolic score 3 | 1.23 | 0.69- 2.20 | 0.481 |
| Diastolic score 4 | 2.32 | 1.25- 4.33 | 0.008 |
| Diastolic score 5 or 6 | 5.32 | 2.44- 11.56 | <0.001 |
| Age at enrollment | 1.73 | 1.25-2.40 | 0.001 |
| Male sex | 1.34 | 0.89- 2.01 | 0.164 |
| Body mass index | 0.93 | 0.76-1.13 | 0.447 |
| Systolic blood pressure | 1.01 | 0.84-1.22 | 0.880 |
| Active smoking | 2.16 | 1.35-3.47 | 0.001 |
| Estimated glomerular filtration rate | 0.97 | 0.78-1.21 | 0.767 |
| HDL cholesterol | 0.78 | 0.62-0.98 | 0.036 |
| Non-HDL cholesterol | 1.03 | 0.86-1.23 | 0.733 |
| Hypertensive medication | 1.19 | 0.75-1.89 | 0.461 |
| Lipid-lowering medication | 1.15 | 0.74-1.80 | 0.534 |
| CVD history | 2.37 | 1.46-3.79 | <0.001 |
| Diabetes mellitus history | 1.69 | 1.05-2.70 | 0.030 |
| Left ventricular hypertrophy | 1.72 | 0.96-3.08 | 0.068 |
| Ejection fraction <50% | 1.00 | 0.38-2.59 | 0.996 |

Hazard ratios of continuous variables were scaled to half the 16^th^-84^th^ percentile range.

CI, confidence interval; CVD, cardiovascular disease; HDL, high-density lipoprotein.

**Supplemental Figure 1. (A) Left ventricular diastolic dysfunction (LVDD) classification per the ASE/EACVI algorithm in the reference group. (B) Age- and sex-stratified distribution of individuals with left ventricular diastolic dysfunction (LVDD) in the reference group.**

*100% of cases in the LVDD group had LVDD Grade Indeterminate.
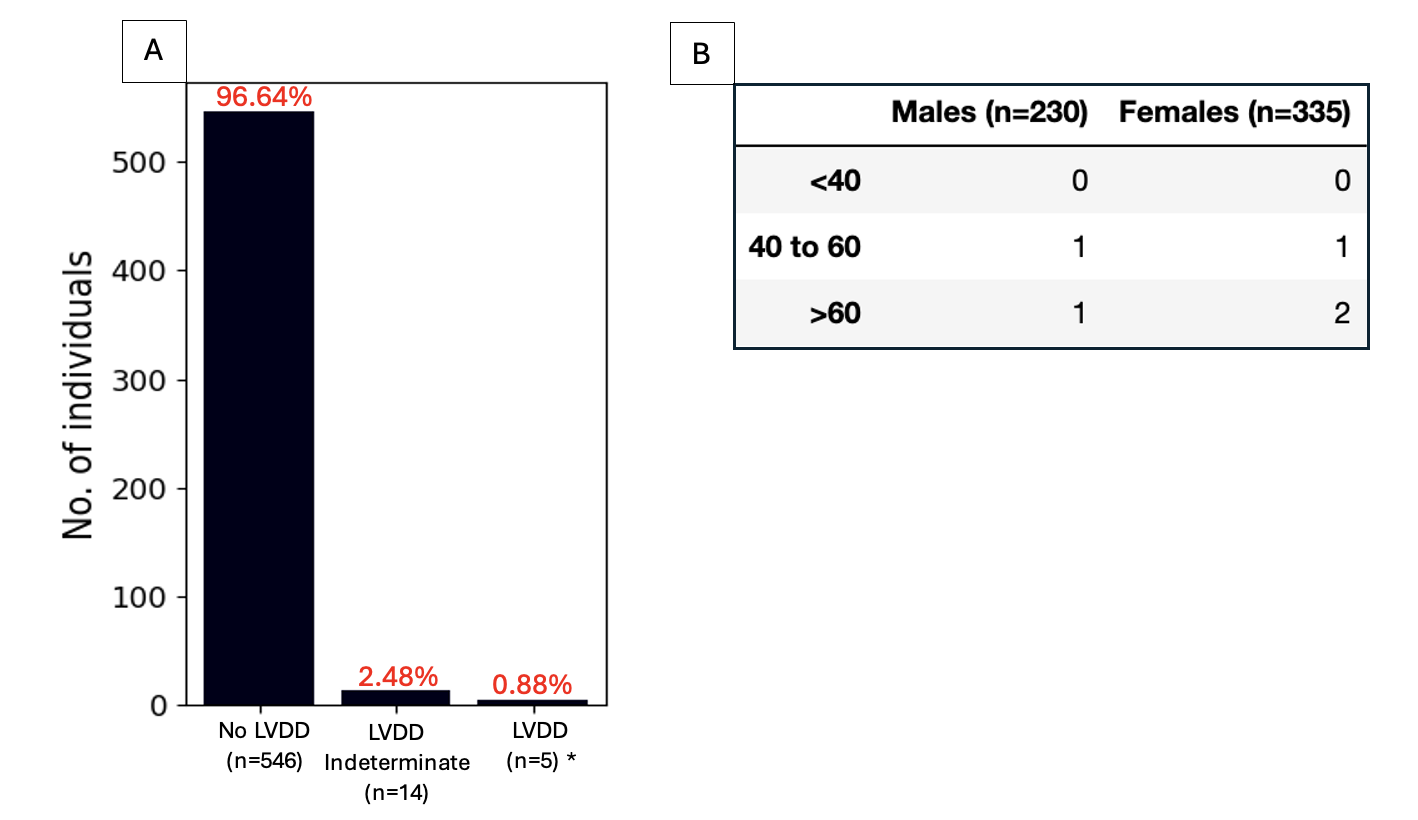

**Supplemental Figure 2. Association of clinical correlates with LVDD using the PBHS diastolic score.** Values represent the odds of having a PBHS diastolic score ≥4. Odds ratios of continuous values were scaled to half the 16th-84th percentile range, which is equal to 19 years for age, 11 mmHg for pulse pressure, 37 mg/dL for non-HDL cholesterol, and 19 mg/dL for HDL cholesterol.

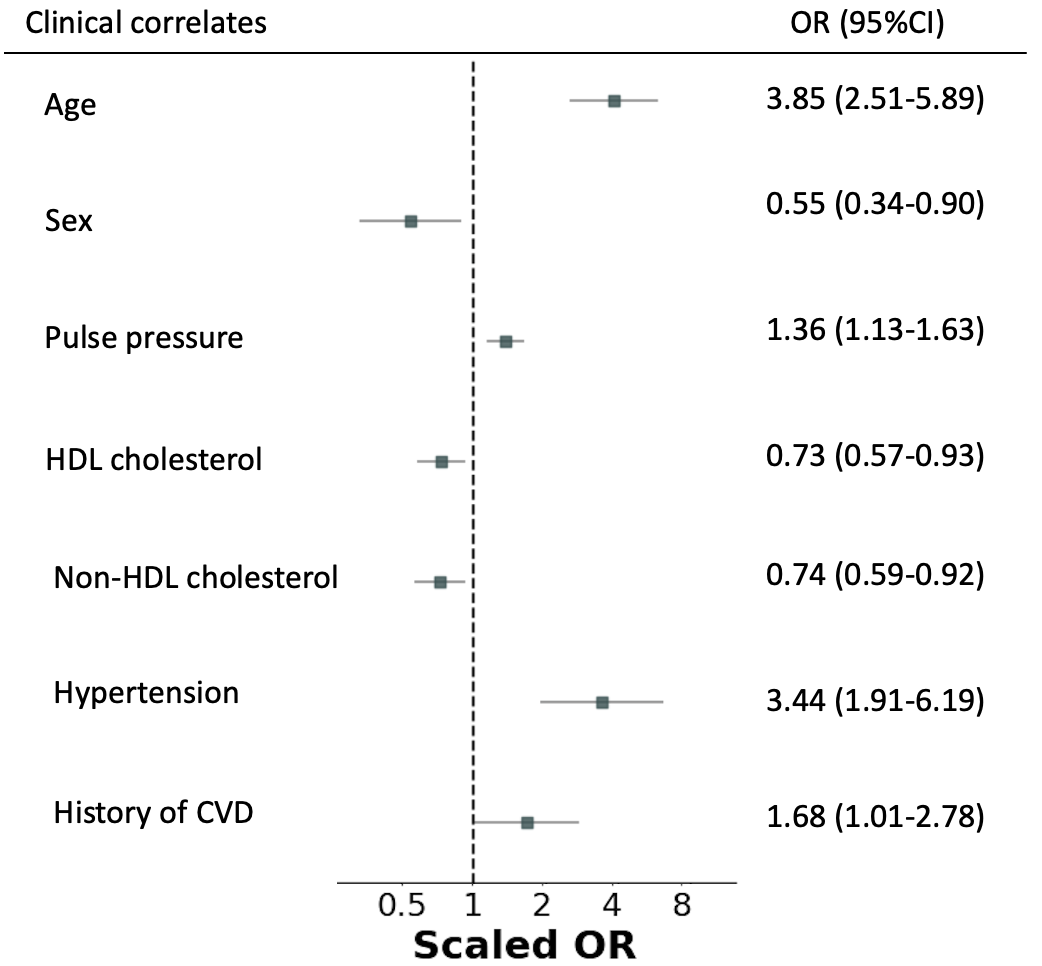

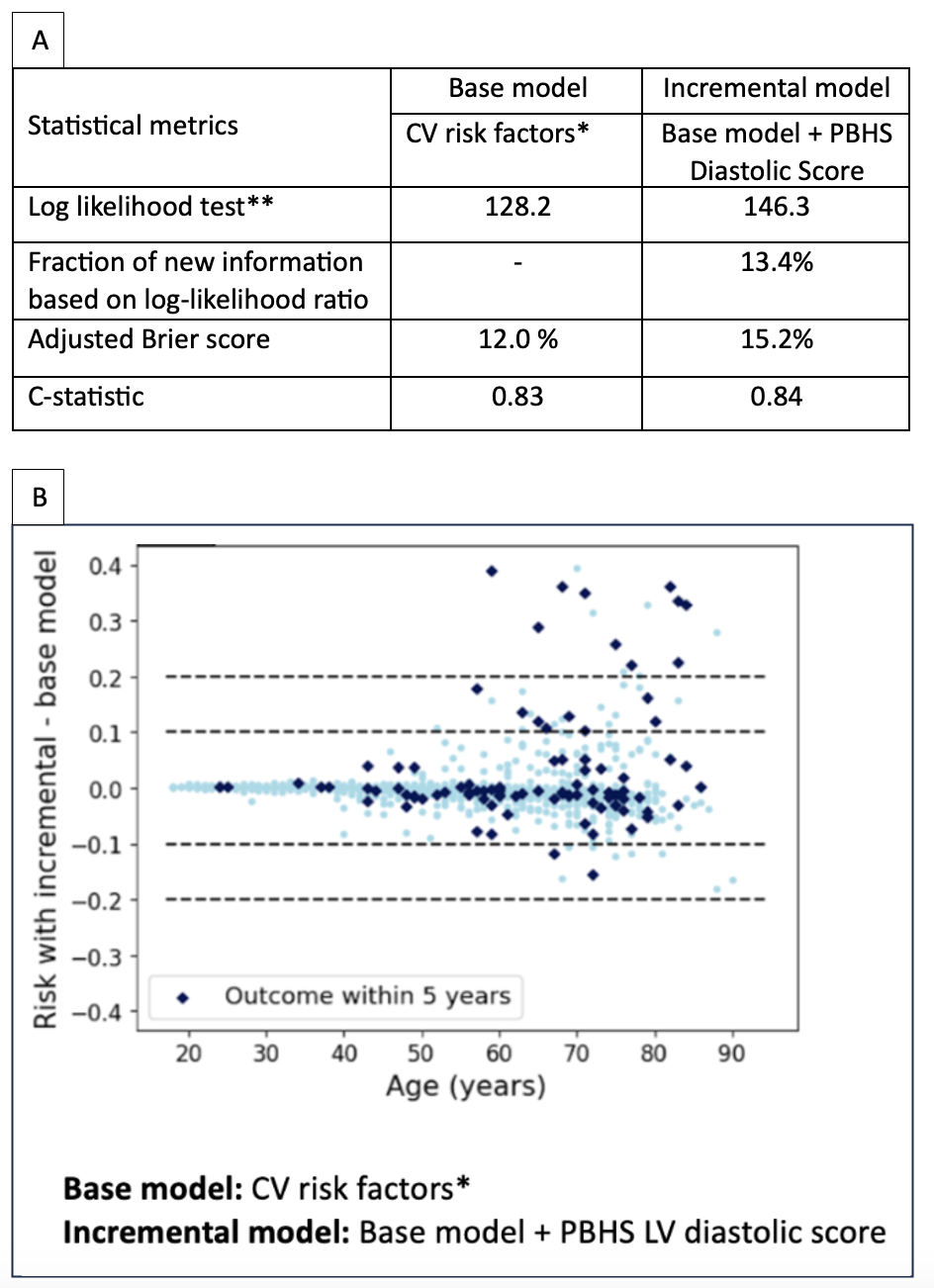
**Supplemental Figure 3. The value of incremental risk models considering LV diastolic function (LVDF) assessment by the PBHS diastolic score in predicting MACE.** (A) Statistical performance metrics of the base model supplemented with LVDF assessment. (B) Scatter plot of the risk difference between the base model and the incremental model with LVDF assessment.

*Base model cardiovascular risk factors include age, sex, body mass index, systolic blood pressure, active smoking status, serum HDL cholesterol, non-HDL cholesterol, estimated glomerular filtration rate, lipid-lowering medication use, hypertensive medication use, history of diabetes mellitus, history of CVD, LVEF <50%, and LV mass index.

**P-value <0.001 for the difference in log likelihood between base and incremental model.
